# Loss-of-function variants in the *KCNQ5* gene are associated with genetic generalized epilepsies

**DOI:** 10.1101/2021.04.20.21255696

**Authors:** Johanna Krüger, Julian Schubert, Josua Kegele, Audrey Labalme, Miaomiao Mao, Jacqueline Heighway, Guiscard Seebohm, Pu Yan, Mahmoud Koko, Kezban Aslan, Hande Caglayan, Bernhard J. Steinhoff, Yvonne G. Weber, Pascale Keo-Kosal, Samuel F. Berkovic, Michael S. Hildebrand, Steven Petrou, Roland Krause, Patrick May, Gaetan Lesca, Snezana Maljevic, Holger Lerche

## Abstract

**Objective:** *De novo* missense variants in *KCNQ5*, encoding the voltage-gated K^+^ channel K_V_7.5, have been described as a cause of developmental and epileptic encephalopathy (DEE) or intellectual disability (ID). We set out to identify disease-related *KCNQ5* variants in genetic generalized epilepsy (GGE) and their underlying mechanisms.

**Methods:** 1292 families with GGE were studied by next-generation sequencing. Whole-cell patch-clamp recordings, biotinylation and phospholipid overlay assays were performed in mammalian cells combined with docking and homology modeling.

**Results:** We identified three deleterious heterozygous missense variants, one truncation and one splice site alteration in five independent families with GGE with predominant absence seizures, two variants were also associated with mild to moderate ID. All three missense variants displayed a strongly decreased current density indicating a loss-of-function (LOF). When mutant channels were co-expressed with wild-type (WT) K_V_7.5 or K_V_7.5 and K_V_7.3 channels, three variants also revealed a significant dominant-negative effect on WT channels. Other gating parameters were unchanged. Biotinylation assays indicated a normal surface expression of the variants. The p.Arg359Cys variant altered PI(4,5)P_2_- interaction, presumably in the non-conducting preopen-closed state.

**Interpretation:** Our study indicates that specific deleterious *KCNQ5* variants are associated with GGE, partially combined with mild to moderate ID. The disease mechanism is a LOF partially with dominant-negative effects through functional, rather than trafficking deficits. LOF of K_V_7.5 channels will reduce the M-current, likely resulting in increased excitability of K_V_7.5- expressing neurons. Further studies on a network level are necessary to understand which circuits are affected and how the variants induce generalized seizures.

## INTRODUCTION

*KCNQ5* is one of five members of the highly conserved *KCNQ* gene family, which encodes the α- subunits of the M-type, voltage-gated delayed rectifier potassium channels K_v_7.1-7.5^1,2^. Four subunits can form a functional channel as either homo- or heterotetramers. Heteromeric channels containing K_v_7.3 subunits together with K_V_7.2 or K_V_7.5 have been shown to yield larger K^+^ currents than each of the subunits alone^1,2,3^. *KCNQ1* is mainly expressed in cardiac muscle and the cochlea^4^, *KCNQ2* and *-3* are mainly expressed in the brain and peripheral nerves^5^, *KCNQ4* is expressed in sensory outer hair cells^6^ and *KCNQ5* is expressed in the brain, skeletal muscle and blood vessels^1,2,7^. Accordingly, K_v_7.1 rather interacts with auxiliary *KCNE* subunits than any of the neuronal K_v_7 subunits, and variants in either one of them can cause cardiac arrhythmia or deafness^4,8^. Due to its very isolated expression, *KCNQ4* is a key contributor to the auditory system and its dysfunction has been associated with non-syndromic dominant deafness^6^.

The other three family members, *KCNQ2, -3* and *-5*, are important regulators of the neuronal M-current within the nervous system that very effectively controls neuronal firing^9^. In particular, dominant negative K_v_7.5 channel expression has been found to decrease the medium and slow afterhyperpolarization currents in the CA3 region of the hippocampus in a mouse model^10^. Moreover, this model unraveled the role of *KCNQ5* in attenuating synaptic inhibition and modifying hippocampal network synchronization^11^. Upon application of muscarinic receptor agonists, such as acetylcholine, a signaling cascade is triggered that causes depletion of phosphatidylinositol-4,5-bisphosphate (PI(4,5)P_2_) from the membrane, which forces K_v_7 channels to close reducing the M-current and resulting in increased neuronal firing^12^.

Loss-of-function (LOF) variants in *KCNQ2* and *KCNQ3* were first identified as the cause of epileptic seizures in patients with benign familial neonatal epilepsy (BFNE) and later in developmental and epileptic encephalopathies (DEE)^13,14,4,15^. More recently, four *de novo* heterozygous missense variants in *KCNQ5* in individuals with intellectual disability (ID) alone or with treatment-resistant DEE have been described. These variants lead to a LOF by depolarizing shifts in activation, slowed activation kinetics or reduced cell surface expression in three individuals, or a gain-of-function (GOF) characterized by a hyperpolarizing shift in activation, accelerated activation kinetics and slowed deactivation kinetics in one of the DEE patients, who displayed the most severe phenotype of all^16^. Additionally, an individual presenting with absence seizures in adolescence, migraine and mild ID has recently been identified with an intragenic duplication of *KCNQ5* most likely causing haploinsufficiency by skipping exons 2-11 and resulting in a premature stop codon^17^.

Here, we analyzed different cohorts of GGE patients to identify causative variants in *KCNQ5*, studied the phenotypes of affected individuals and co-segregation of the detected variants. Electrophysiological and biochemical characterization of variants contributing to the disease was performed by expression of mutant K_v_7.5 subunits in Chinese hamster ovary (CHO) cells and using whole cell patch-clamping, biotinylation and phospholipid overlay assays.

## SUBJECTS/MATERIALS AND METHODS

### Study participants

This study was approved by the local institutional review boards of the participating centers. The patients or their relatives gave their written informed consent. In total, 12 individuals from 7 families were ascertained from three different cohorts. Individuals 1, 2, 3, 4, 6 and 7 were ascertained from the EuroEPINOMICS-CoGIE study, while individual 5 was ascertained from the Epi25 study (Tuebingen subcohort), individuals 8 and 9 were analyzed in a gene panel, and individual 10 and two additional individuals were ascertained from the EPGP/Epi4K study. Medical and family histories, neurological examination, brain imaging and EEG findings were analyzed. Seizure types were classified according to the latest International League Against Epilepsy classification^18^. Blood samples were taken from available family members and DNA was extracted by standard procedures.

### Genetic Analysis

Individuals from all cohorts were whole exome sequenced, whereas individuals 8 and 9 were analyzed in an epilepsy gene panel. Validation of discovered *KCNQ5* variants was performed via Sanger sequencing with the exception of family 3 due to insufficient amount of DNA. Splice site variants were further investigated by RNA extraction from patient blood (PAXgene Blood RNA System, BD), and subsequent cDNA amplification and sequencing.

### Variant Interpretation

*KCNQ5* variants were considered putatively disease-relevant if 3/4 of the following criteria were met (1) likely functional effect (protein-truncating variant, inframe-deletion or a missense variant with at least 4 pathogenic predictions), (2) MAF of 0 in the European populations of the 1000 genomes (http://www.1000genomes.org), the Exome Variant Server (EVS; http://evs.gs.washington.edu) and in the Genome Aggregation Database (gnomAD; http://gnomad.broadinstitute.org), (3) confirmed in all affected family members, and (4) the variant demonstrated an abnormal effect in electrophysiological recordings or analysis of splicing. The variants were interpreted according to the American College of Medical Genetics and Genomics standards and guidelines for the interpretation of sequence variants^19^.

### Testing for variant enrichment

To estimate the burden of qualifying variants in *KCNQ5* in GGE cases vs. controls, we extended the rare-variant association analysis performed by Epi4K/EPGP^20^ using additional cohorts of unrelated GGE patients and matched controls, all of European ancestry. A combined non-overlapping set of 4,418 GGE cases and 7,727 controls from these previously published cohorts was investigated using the Cochran-Mantel-Haenszel exact test as follows: 1) 640 cases and 3,877 controls from the Epi4K/EPGP study^20^, 2) 874 cases and 2,177 matched controls from EuroEPINOMICS-CoGIE, EpiPGX & CENet consortia studies^21,22^, 3) 2,904 cases and 1,763 matched controls from the Epi25 Collaborative study^23^. Qualifying variants were defined consistently across cohorts using the criteria previously defined by Epi4K/EPGP^20^.

### Functional analysis

#### Mutagenesis

pcDNA3.1-P2A-eGFP and pcDNA3.1-P2A-tagRFP vectors containing the human K_v_7.5-subunit cDNA (NM_019842.3) were purchased from GeneScript (Netherlands). Site-directed mutagenesis was performed using PCR with Pfu polymerase (Promega, Germany). The inserts were sequenced to confirm the introduction of the point mutations and to exclude additional alterations.

#### Transfection and expression in CHO cells

CHO-K1 cells were cultured at 37°C in a 5% CO_2_ humidified atmosphere and grown in Ham’s F12 containing 2mM glutamine (Gibco), 10% (v/v) fetal calf serum (PAN Biotech) and 1% Penicillin-Streptomycin (Gibco; only in the Australian cohort). Transfection was performed 24-48h prior to electrophysiological recordings with Lipofectamine 3000 (Invitrogen) following the manufacturer’s protocol using 2-2.5µg DNA and an additional 0.25µg of eGFP DNA for the truncation variant (p.Ala301Glyfs*64) lacking the fluorescent marker due to the early stop codon. For co-transfection with WT subunits the same protocol was applied using 2-2.5µg of DNA in total in a molar ratio of 1:1 and only 1µg DNA for the WT controls. For co-expression of K_v_7.3, a CHO-K1 line stably expressing K_V_7.3 channels was used and transiently transfected as mentioned above. This cell line was created by Lenti-viral transduction of CHOs using a pLenti4/TO/V5-DEST gateway vector carrying human K_v_7.3 WT cDNA. Zeocin was used for selection of transduced cells and removed 48h prior to recordings. Expression of the introduced K_v_7.3 WT was further confirmed by Western blot.

#### Electrophysiology

Standard whole-cell patch clamp recordings were performed using an Axopatch 200B or Multiclamp 700B amplifier, a Digidata 1320A, 1440A or 1550B digitizer (Axon Instruments), and pCLAMP 8, 10.4 or 11.1 data acquisition software (Molecular Devices). Leakage and capacitive currents were automatically subtracted using a pre-pulse protocol (-P/4). Cells were held at -80mV in whole-cell configuration for 2min prior to recording and series resistance was compensated (at approx. 85%) and monitored regularly. Currents were filtered at 1 kHz and digitized at 5 kHz. The bath solution contained (in mM): 138 NaCl, 2 CaCl2, 5.4 KCl, 1 MgCl2, 10 glucose and 10 (4-(2-hydroxyethyl)-1-piperazineethanesulphonic acid (HEPES) (pH 7.4 adjusted with NaOH). Borosilicate glass pipettes had a final tip resistance of 1.5-3.5MΩ and were filled with pipette solution containing (in mM): 140 KCl, 2 MgCl2, 10 EGTA, 10 HEPES, 5 K_2_ATP (pH 7.4 adjusted with KOH)^24^. All recordings were performed at room temperature of 21-23°C (RT).

Cells were visualized using an inverted microscope (Axio-Vert.A1, Zeiss or Nikon Eclipse). In case of single plasmid transfections only green (eGFP) or red (tagRFP) fluorescent cells were selected for electrophysiological recordings 24-48h after transfection, whereas in co-transfection recordings, cells were selected that showed an approximately equal amount of both, red and green fluorescence.

#### Patch clamp protocols and data analysis

K^+^ currents were induced by depolarizing the membrane from a holding potential of -80mV to +60mV in 10mV steps for 2s. Subsequently, a shorter hyperpolarizing pulse was elicited to -120mV for 0.5s to obtain tail currents. Current amplitudes were calculated from the mean steady-state current for the last 0.5s of the first step depolarization. Current densities (pA/pF) were obtained by normalizing the current amplitudes to the cell membrane capacitance. The activation curve was determined by plotting the normalized tail (I_tail_) current against the step potential (V_s_). A Boltzmann function, ***I***_tail_ = 1/(1 + exp[(***V***_0.5_ – ***V***_s_)/***k***]), where ***V***_0.5_ is the voltage of half-maximal activation and ***k*** is the slope factor, was fit to the data points.

Clampfit software of pClamp10.7 (Axon Instruments), Microsoft Excel (Microsoft Corporation, Redmond, WA, USA) or GraphPad software (GraphPad Prism 8, San Diego, CA, USA) were used for data and statistical analysis. All data is shown as mean ± SEM. All data were tested for normal distribution. One-way ANOVA with Dunnett’s *post hoc* test was used to evaluate statistical significance of normally distributed data. If the data was not normally distributed, a Kruskal-Wallis test was performed followed by a Benjamini, Krieger, and Yekutieli test. For all statistical tests p<0.05 was considered significant. Scatter plots show single cell values, median and interquartile range.

#### Western blot analysis

CHO cells were lysed 24h after transfection with either wildtype *KCNQ5*, one of the mutant cDNAs or water (mock) using the following buffer (in mM): 20 Tris (pH 7.5), 150 NaCl, 1 EDTA, 1 EGTA, 2.5 Napyrophosphate, 1 β-glycerolphosphate, 1 sodium-orthovanadate, 10 DTT, 1% Triton and 1x cOmplete solution (Roche). Total protein concentration was measured via Bradford assay. 8% polyacrylamid gels were loaded with 20µg of total protein per lane to separate these by sodium dodecyl sulfate-polyacrylamide gel electrophoresis (SDS PAGE). After transferring the proteins onto a nitrocellulose membrane (Whatman) via electrophoresis at 4°C in Towbin buffer (25 mM Tris, 192 mM glycine, pH 8.3, 10% (v/v) methanol), the blots were blocked in 5% non-fat dry milk powder in phosphate-bufferd saline with 1% Tween (PBST) for 1h at RT. Subsequently, membranes were probed with a polyclonal rabbit primary antibody against K_v_7.5 (ABN1372, Millipore) at 1:7500 and a monoclonal mouse primary antibody against vinculin (V9131, Sigma-Aldrich) at 1:5000 overnight at 4°C. Following this, the membranes were washed and shaken in PBST thrice and then re-probed with a secondary goat anti-rabbit IgG-HRP-conjugated antibody (172-1019, Bio-Rad) at 1:10000 or secondary goat anti-mouse IgG-HRP-conjugated antibody (172-1011, Bio-Rad) at 1:10000, respectively, for 1h at RT. After three more washing steps in PBST, detection was performed via enhanced chemiluminescence (ECL; Amersham, Cytiva).

#### Biotinylation Assay

For isolation of membrane proteins, the Pierce Cell Surface Protein Biotinylation and Isolation Kit (ThermoFisher) was used according to the manufacturer’s protocol. In brief, cells were cultured for 48h and transfected with either the WT or one of the mutant plasmids. Cells were biotinylated, lysed and isolated by binding to probed agarose beads 24h after transfection. After elution, the proteins were prepared for Western blot analysis. Western blots were performed as described above. A polyclonal rabbit anti-actin primary antibody (1:1000, A2066, Sigma-Aldrich) was used as control.

#### Protein-Phospholipid Overlay Assay

PIP strips (Molecular Probes) were used as described in the manufacturer’s protocol. Concisely, cells were cultured and transfected as described above and lysed 24h after transfection. PIP strips were blocked in 3% fatty-acid free bovine serum albumin (BSA) in Tris-bufferd saline with 1% Tween (TBST) for 1h at RT. Next, membranes were incubated at 4°C overnight in TBST+3% BSA and a final protein concentration of 1µg/ml. Membranes were washed and Western blot was performed as described above.

#### Docking, homology modeling, and molecular dynamics simulations

Molecular dynamics simulations were conducted as described before^25^. In brief, human K_V_7.5 models were constructed for K_V_7.5, var1 (UniProtKB - Q9NR82 (KCNQ5_HUMAN)). Homology modeling, docking experiments, and molecular dynamics simulations were performed using YASARA Structure, version 20.10.4. Models were constructed based on published Cryo-EM and crystal structures with PDB numbers *6V01* (closed channel model) and *2nz0, 3rbz, 5vms* (open channel model)^26,27,28,29,30^. PI(4,5)P_2_ was positioned analogous to template *6V01*.*pdb* (K_V_7.5-closed state model) or docked with PI(4,5)P_2_ head groups in proximity to the S4-S5 linker α-helices (K_V_7.5-open state model)^31^. The resultant models were mutated at all p.Arg359 positions to cysteines resulting in K_V_7.5- p.Arg359Cys mutant channel models. All models were energy minimized in phosphatidyl-ethanolamine (1-palmitoyl, 2-oleoyl) membranes and all atoms free MD simulations started for the indicated times. MD simulation settings were: pH=7.4, ion concentration (mass fraction) were 0.9% NaCl (physiological solution), ForceField AMBER14, temperature=‘298K’, electrostatics set to cutoff=8, pressure pressure=1 bar.

## RESULTS

### GENETIC SCREENING OF GGE COHORTS

In total, we identified 10 individuals within 5 families with a GGE phenotype carrying a potentially disease-associated variant (see criteria in methods) in *KCNQ5* (depicted in Fig 1A). p.Arg359Cys (1 family with 4 affected members in 2 generations, all carrying the variant, phenotypes: 1 CAE, 1 IGE + mild ID, 1CAE, JME + moderate ID, 1 CAE + mild developmental delay, 1 asymptomatic carrier) and p.Gln735Arg (2 JME, 1 asymptomatic carrier) were first detected, each in 1 of 238 independent GGE families from the EuroEPINOMICS-CoGIE cohort^21^. Both variants co-segregated in 2-4 affected family members and were found each also in one asymptomatic carrier. This prompted us to search for further variants in three other cohorts. p.Leu692Val (CAE, IGE) was found in 1 individual of 339 GGE families in our Epi25 subcohort, p.Glu265_Thr306del in 2 related individuals (CAE + mild ID, CAE, IGE + mild ID) in an epilepsy gene panel that was performed in 75 individuals, and p.Ala301Glyfs*64, in 1 individual (JME) of 640 GGE families from the Epi4K/EPGP cohort^20^. Two additional variants from the Epi4K/EPGP cohort, p.Phe165Ile and p.Leu926Ser, were found each one time in the gnomAD or TOPMed databases and did not show any alterations in electrophysiological recordings as compared to the WT (see Table 2), hence, the individuals carrying these benign variants will not be considered further on.

Pedigrees and detailed clinical descriptions are available upon request.

**Figure 1.**
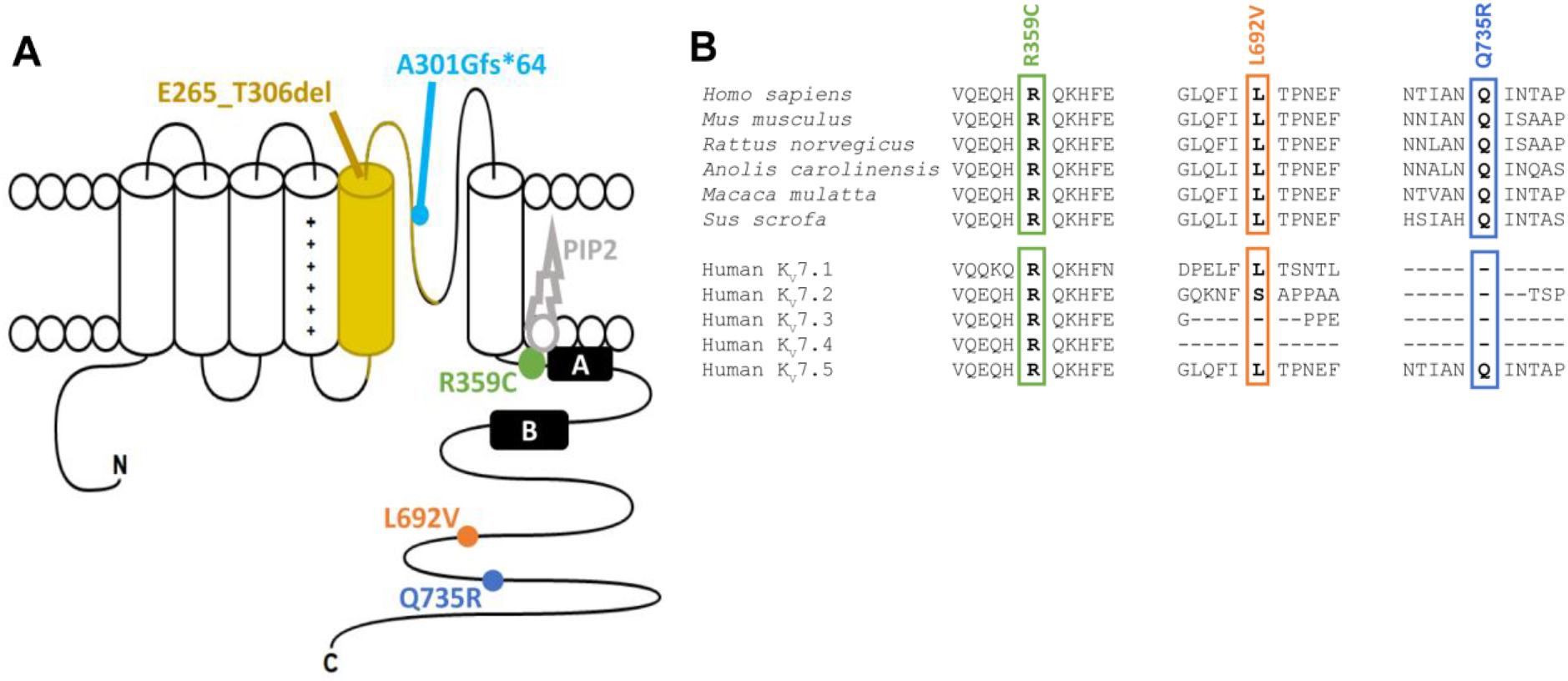
Variants affecting the K_v_7.5 potassium channel. (**A**) Schematic of the K_v_7.5 subunit of which four assemble to form a channel. Each subunit consists of a voltage-sensor domain (S1-S4) and a pore forming region (S5 and S6) including the pore loop. Point mutations are localized in highly conserved regions of the C-terminus (R359C, L692V, Q735R), while a splice site variant causes a deletion of the S5 segment and parts of the pore forming loop (E265_T306del) and a frame-shift mutation leads to an early stop codon in the pore loop (A301Gfs*64). (**B**) Amino acid alignment across multiple species (top) and across all human K_V_7 family members (bottom) shows evolutionary conservation of R359, L692, Q735 and their surrounding amino acids in K_V_7.5. Species from top to bottom: human, mouse, rat, Carolina anole, macaque, pig.

Statistical analysis was performed to validate *KCNQ5* as a potentially predisposing gene for GGEs. The odds of the occurrence of qualifying variants (as previously defined by the Epi4K/EPGP^20^) in *KCNQ5* in GGE cases vs. controls was calculated over three large previously published cohorts (partially overlapping with the discovery cohorts mentioned above), each matched with independent controls. In total, we identified 8 qualifying variants in cases vs. 3 in controls, with a stratified odds ratio of 10.6 (confidence interval: 2.4-64.8; *p* = 0.00044). The odds were homogeneous across cohorts (Breslow-Day test; *p* = 0.9). This suggests that *KCNQ5* variants are enriched in GGE cases, considering this hypothesis driven approach.

### FUNCTIONAL CHARACTERIZATION OF *KCNQ5* VARIANTS

The *KCNQ5* c.918+5G>A (p.Glu265_Thr306del) variant in individuals 8 and 9 affects a donor splice-site. Splicing prediction tools predicted abolition of the donor site (SplceAI score 0.98). To examine the consequences of this variant, RNA was extracted from blood, and analysis of cDNA amplicons with primers located in exon 2 and at the junction of exon 6 and 7 was performed and revealed the expected 583-bp band for the wild type (WT), and an additional smaller band of 460 bp, corresponding to the complete skipping of exon 5. This was confirmed by sequencing of the PCR products. This band was absent from the cDNA of the controls. To this end, the variant was assumed a LOF due to the large deletion within the critical pore region of the channel (S5 segment; see Fig 1A).

To functionally characterize the remaining variants, CHO cells were transfected either with the mutant *KCNQ5* cDNA and compared to those transfected with WT *KCNQ5* cDNA (2-2.5µg), or with a 1:1 mix of mutant and WT cDNA (1µg each) and compared to the same amount of WT cDNA alone (1µg) to test for a dominant-negative effect of mutant on WT channels. Standard whole-cell patch-clamp recordings were performed from transfected cells which were identified via fluorescent markers. Figure 2A displays representative raw current traces recorded from cells expressing either WT or one of the mutant channel subunits. Untransfected cells were used as additional controls. Homomeric expression of all three mutated channel subunits caused a significant reduction in peak current amplitude and current density (Fig 2B). The p.Arg359Cys variant presented the most severe reduction being almost indistinguishable from untransfected control cells (peaks at 25.9 ± 5.9pA/pF and 15.5 ± 8.4pA/pF, respectively; both *n = 10*; see Table 1). p.Leu692Val and p.Gln735Arg reached comparable current densities (112.3 ± 32.2pA/pF and 139.6 ± 21.4pA/pF, respectively; both *n = 10*), of about 20% of the WT (620.9 ± 133.3pA/pF; *n = 11*; Fig 2C). The voltage dependence of activation, as derived from normalized tail currents, was not changed for p.Leu692Val and p.Gln735Arg, and could not be evaluated for p.Arg359Cys, since this variant exhibited too small currents (Fig 2D and Table 1).

**Table 1.** Biophysical properties of K_v_7.5 WT and variant channels (Hertie Institute for Clinical Brain Research, Tübingen, Germany).

|  | Homomeric expression |  |  |  |  | Heteromeric expression |  |  |  |  | Heteromeric expression + KCNQ3-WT |  |  |  |  |
| --- | --- | --- | --- | --- | --- | --- | --- | --- | --- | --- | --- | --- | --- | --- | --- |
|  |  |  | Activation kinetics |  |  |  |  | Activation kinetics |  |  |  |  | Activation kinetics |  |  |
|  | Current density [pA/pF] | n | V<sub>1/2</sub> [mV] | k | n | Current density [pA/pF] | n | V<sub>1/2</sub> [mV] | k | n | Current density [pA/pF] | n | V<sub>1/2</sub> [mV] | k | n |
| **WT** | 620.9 ± 133.3 | 11 | -29.91 ± 5.26 | -17.31 ± 1.93 | 11 | 487.8 ± 54.6 | 12 | -26.40 ± 3.16 | -19.26 ± 3.32 | 12 | 451.1 ± 44.0 | 13 | -24.05 ± 3.17 | -17.60 ± 2.13 | 13 |
| **R359C** | 25.9 ± 5.9<sup>a</sup> | 10 | - | - | - | 49.1 ± 10.7<sup>a</sup> | 10 | - | - | - | 106.8 ± 32.5<sup>c</sup> | 10 | -28.68 ± 3.58 | -8.80 ± 0.56 | 7 |
| **L692V** | 112.3 ± 32.2<sup>a</sup> | 10 | -29.51 ± 2.66 | -11.98 ± 1.67 | 9 | 210.7 ± 43.5<sup>b</sup> | 10 | -24.76 ± 3.29 | -18.22 ± 2.57 | 10 | 171.5 ± 31.5<sup>d</sup> | 10 | -18.45 ± 5.97 | -15.43 ± 2.33 | 9 |
| **Q735R** | 139.6 ± 21.4<sup>a</sup> | 10 | -27.35 ± 6.41 | -15.95 ± 3.91 | 9 | 217.5 ± 61.9<sup>b</sup> | 10 | -19.98 ± 5.45 | -17.75 ± 1.56 | 9 | 226.3 ± 66.7<sup>d</sup> | 10 | -26.40 ± 2.28 | -15.19 ± 2.00 | 9 |

<sup>a</sup> $p < 0.0001$ via one-way ANOVA with post hoc correction for multiple comparisons with Dunnett's test.
<sup>b</sup> $p < 0.001$ via one-way ANOVA with post hoc correction for multiple comparisons with Dunnett's test.
<sup>c</sup> $p < 0.0001$ via Kruskal-Wallis test with post hoc correction for multiple comparisons with Benjamini, Krieger and
Yekutieli's test.
<sup>d</sup> $p < 0.05$ via Kruskal-Wallis test with post hoc correction for multiple comparisons with Benjamini, Krieger and

**Figure 2.**
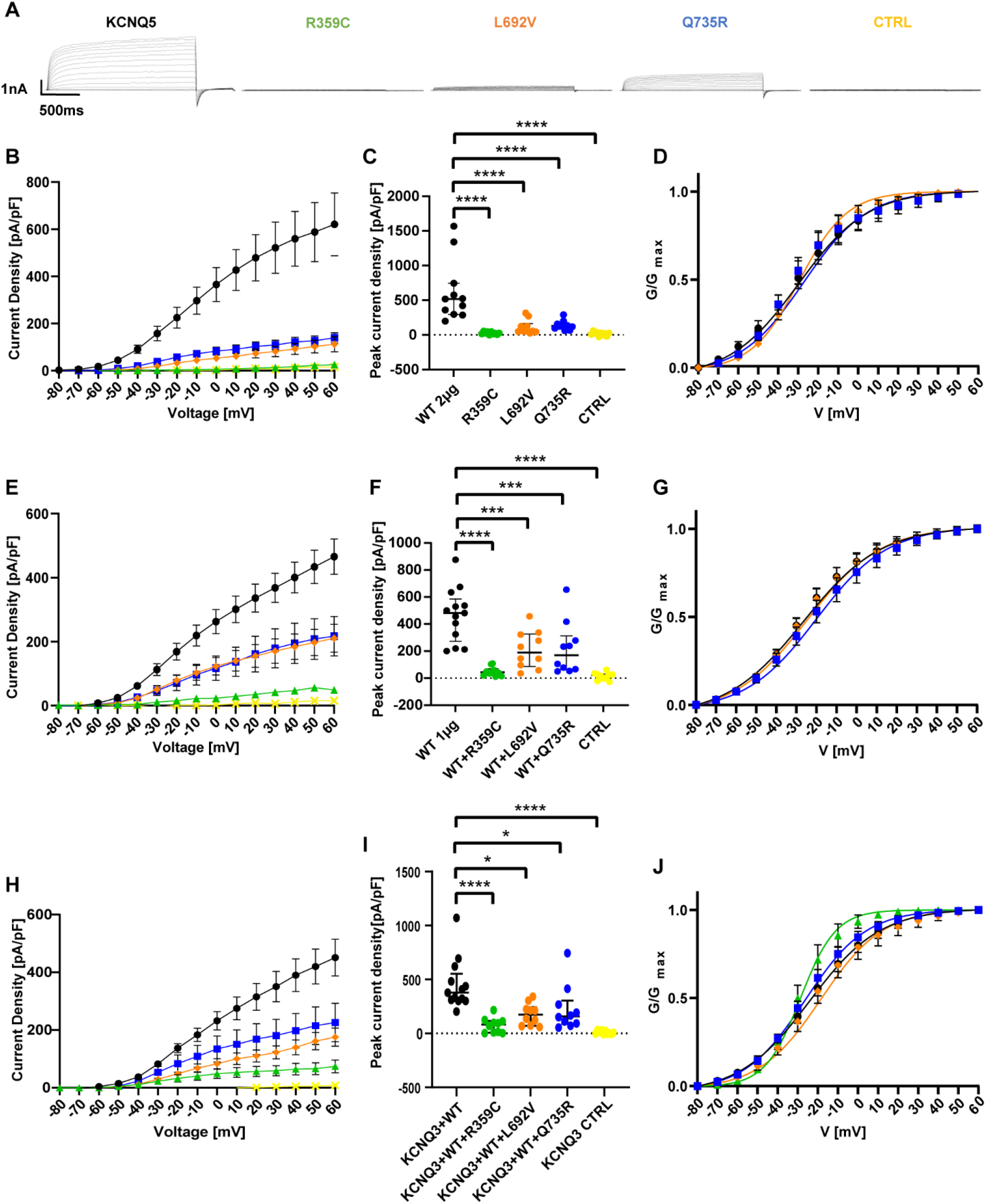
Functional effects of K_v_7.5 WT and mutant channels in Chinese hamster ovarian cells. (**A**) Representative K^+^ current traces from KCNQ5 WT (black), R359C (green), L692V (orange), Q735R (blue) and untransfected control cells (CTRL, yellow) during voltage steps from -80 mV to +60 mV in 10 mV increments. (**B**) Peak K+ currents of cells either transfected with WT or one mutated channel subunit were normalized by cell capacitances and plotted versus voltage. All variants result in a significant reduction in current density compared to the WT. WT, n = 12; WT + R359C, n = 10; WT + L692V, n = 10; WT + Q735R, n = 10; CTRL, n = 10. (**C**) Comparison of maximum peak current density at +60 mV. All variants show a significant reduction compared to the WT. (**D**) Voltage-dependent activation curves. Lines represent Boltzmann functions fit to the normalized tail current. Currents in the R359C variant were too small to establish such a relationship. (**E**) Peak K+ currents normalized by cell capacitances and plotted versus voltage of cells either transfected with WT (1µg) or WT and one mutated channel subunit (1µg + 1µg). The significant reduction persisted in all variants compared to the WT indicating a dominant negative effect of the variants on the WT. WT, n = 11; R359C, n = 10; L692V, n = 10; Q735R, n = 10; CTRL, n = 10. (**F**) Comparison of maximum peak current density at +60 mV. All variants show a significant reduction compared to the WT. (**G**) Voltage-dependent activation curves. Lines represent Boltzmann functions fit to the normalized tail current. Currents of the WT + R359C were still too small to establish such a relationship. (**H**) Peak K+ currents normalized by cell capacitances and plotted versus voltage of cells either transfected with WT (1µg) or WT and one mutated channel subunit (1µg + 1µg) in a CHO line stably transfected with KCNQ3-WT. The dominant negative effect of the variants on the WT and the significant reduction persisted in all variants compared to the WT. WT, n = 13; R359C, n = 10; L692V, n = 10; Q735R, n = 10; CTRL, n = 10. (**I**) Comparison of maximum peak current density at +60 mV. All variants show a significant reduction compared to the WT. (**J**) Voltage-dependent activation curves. Lines represent Boltzmann functions fit to the normalized tail current. Shown are means ± SEM (**B, D, E, G, H, J**). Scatter-and-whisker plots (**C, F, I**) show median (horizontal line) and the interquartile ranges. Dots indicate maximum values of single cells. *p ≤ 0.05; ** p≤ 0.01; *** p≤ 0.001; **** p≤ 0.0001; **Table 1** provides exact values and statistical analyses.

Heteromeric expression of mutant and WT subunits in a 1:1 ratio did increase the current density for all variants compared to homomeric expression (Fig 2E). We observed a dominant negative effect for all three variants which was most severe for p.Arg359Cys (peak current density of 49.1 ± 10.7pA/pF, *n = 10*) corresponding to 10% of the WT amplitude (487.8 ± 54.6pA/pF; *n = 12*), while p.Leu692Val and p.Gln735Arg reached less than 45% of the WT (210.7 ± 43.5pA/pF and 217.5 ± 61.9pA/pF, respectively; both *n = 10*) (Fig 2F). The voltage dependence of channel activation was again similar for p.Leu692Val/WT, p.Gln735Arg/WT and WT alone, while p.Arg359Cys/WT-associated amplitudes were still too small for data evaluation (Fig 2G).

Since K_v_7.5 and K_v_7.3 subunits can form heterotetramers^1,2^, heteromeric co-expression of K_V_7.5 WT and mutant subunits in a 1:1 ratio (1µg of each clone) was performed in a CHO cell line stably expressing K_V_7.3 WT channels. Current densities were still significantly reduced for all three investigated variants indicating dominant-negative effects also under these conditions. Peaks reached 24% of the WT (451.1 ± 44.0pA/pF; *n = 13*) for p.Arg359Cys (106.8 ± 32.5pA/pF; *n = 10*), 38% for p.Leu692Val (171.5 ± 31.5pA/pF; *n = 10*), and 50% for p.Gln735Arg (226.3 ± 66.7pA/pF; *n = 10*; Fig 2H and I). Activation curves of all three variants did not significantly differ from the WT (Fig 2J).

In a second series of experiments, the effect of the truncating variant p.Ala301Glyfs*64 and two additional missense variants (p.Phe165Ile and p.Leu926Ser) were investigated. Homomeric expression caused a complete LOF (current density 8.35 ± 1.91pA/pF; *n = 16;* Fig 3A and B) compared to the WT (100.9 ± 7.65pA/pF; *n = 69*), similar to the effect of the p.Arg359Cys. When co-expressed with WT subunits, neither current density nor activation curves were significantly different in cells expressing the WT subunit alone versus cells expressing WT and variant (see Fig 3C to E and Table 2). Both additional missense variants did not show a significant difference in either current density or gating parameters as compared to the WT (see Table 2).

**Figure 3.**
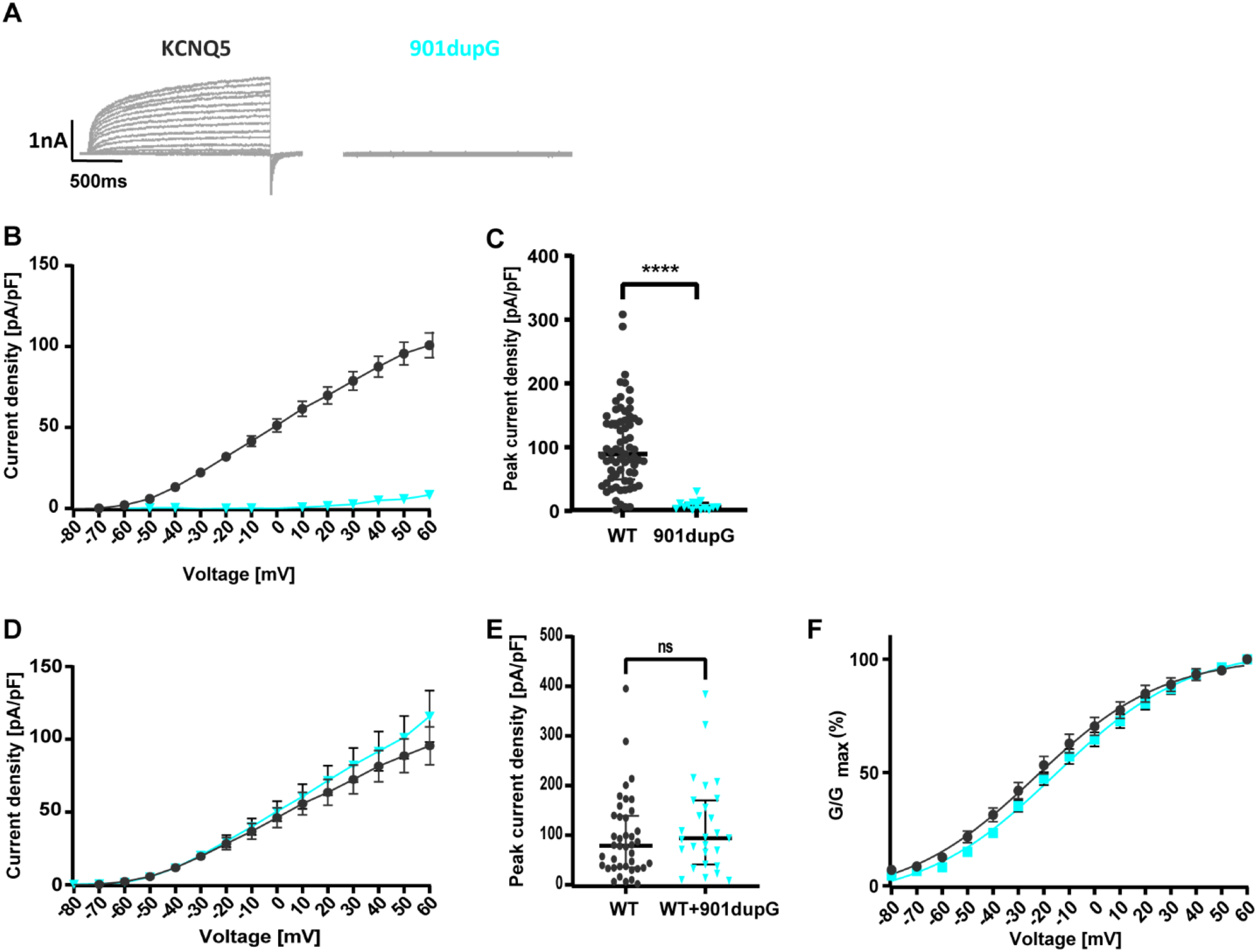
Functional effects of K_v_7.5 WT and mutant channels in Chinese hamster ovarian cells (Australian cohort). (**A**) Representative K^+^ current traces from KCNQ5 WT (black) and c.901dupG (turquoise) during voltage steps from -80 mV to +60 mV in 10 mV increments. (**B**) Peak K+ currents of cells either transfected with WT or c.901dupG channel subunit were normalized by cell capacitances and plotted versus voltage. WT, n = 70; 901dupG, n = 16. (**C**) Comparison of maximum peak current density at +60 mV. c.901dupG results in a significant decrease in current density compared to the WT. (**D**) Peak K+ currents normalized by cell capacitances and plotted versus voltage of cells either transfected with WT (2.5µg) or WT and 901dupG (2.5µg + 2.5µg). No significant differences were observed. WT, n = 40; WT+901dupG, n = 27. (E) Comparison of maximum peak current density at +60 mV. No significant differences were observed. (F) Voltage-dependent activation curves. Lines represent Boltzmann functions fit to the normalized tail current. Shown are means ± SEM (**B, D**,, **F**). Scatter-and-whisker plots (**C** and **E**) show median (horizontal line) and the interquartile ranges. Dots indicate maximum values of single cells. **** p≤ 0.001; ns non significant; **Table 2** provides exact values and statistical analyses.

**Table 2.** Biophysical properties of K_v_7.5 WT and truncated subunits (Florey Institute, Melbourne, Australia).

|  | Homomeric expression |  |  |  |  | Heteromeric expression |  |  |  |  |
| --- | --- | --- | --- | --- | --- | --- | --- | --- | --- | --- |
|  |  |  | Activation kinetics |  |  |  |  | Activation kinetics |  |  |
|  | Current density [pA/pF] | n | V<sub>1/2</sub> [mV] | k | n | Current density [pA/pF] | n | V<sub>1/2</sub> [mV] | k | N |
| **WT** | 100.9 ± 7.65 | 69 | -22.74 ± 2.98 | -20.0 ± 1.83 | 34 | 95.57 ± 12.96 | 40 | -17.29 ± 4.13 | -26.57 ± 2.93 | 24 |
| **901dup G** | 8.35 ± 1.91<sup>a</sup> | 16 | - | - | - | 115.9 ± 17.7 | 27 | -11.73 ± 3.99 | -25.33 ± 1.64 | 23 |
| **F165I** | 153.6 ± 25.48 | 19 | -16.25 ± 3.34 | -21.02 ± 2.31 | 19 | - | - | - | - | - |
| **L926S** | 106.1 ± 7.74 | 26 | -18.96 ± 3.77 | -20.31 | 23 | - | - | - | - | - |

<sup>a</sup> $p < 0.0001$ via Kruskal-Wallis test with post hoc correction for multiple comparisons with Benjamini, Krieger and
Yekutieli's test.

In summary, two missense variants were found to have no functional effect and were thus considered benign, while three other missense variants cause a dominant-negative LOF effect by reducing current density in all three expression conditions, the p.Arg359Cys being the most severe one. The truncated variant (p.Ala301Glyfs*64) has no effect on the WT subunits, only causing a haploinsufficiency. The voltage dependence of channel activation was not significantly changed for any of the variants (Tables 1 and 2).

### PROTEIN PRODUCTION AND MEMBRANE EXPRESSION OF *KCNQ5* VARIANTS IN CHO CELLS

To investigate whether the LOF was caused by a dysfunction in channel opening, or by a trafficking or other defect, the amount of produced protein of the p.Arg359Cys, p.Leu692Val and p.Gln735Arg variants compared to the WT was determined in CHO cells via Western blot. The p.Ala301Glyfs*64 variant had to be excluded from this approach due to the missing C-terminus, which carries the antibody epitope. When whole cell lysates were blotted, no significant difference in protein amount was detected as compared to the WT (Fig 4A). To further examine if the mutant subunits were integrated in the cell membrane, a cell surface protein biotinylation assay with subsequent Western blot was performed. Again, no significant difference between the WT and the variants was observed (Fig 4B). Consequently, the variants do neither alter overall nor cell surface expression of any of the mutant subunits, which suggests that the LOF is likely caused by dysfunctional channel opening and not by a defect in protein production, folding or trafficking.

**Figure 4.**
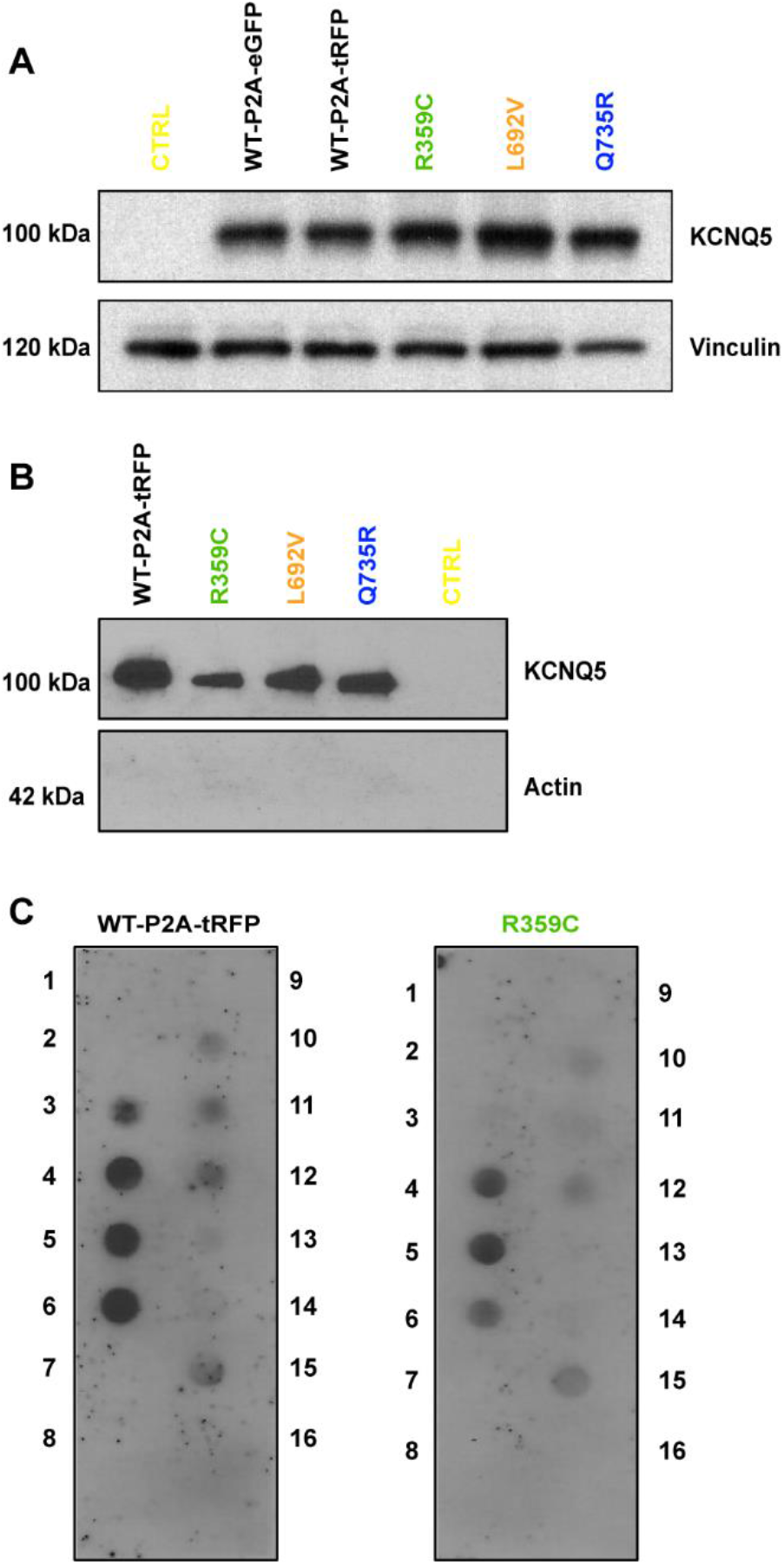
Western blot analysis of *KCNQ5* expression and phospholipid binding abilities in CHO cells. (**A**) Western blot of CHO cell lysates of transiently transfected cells (20µg per lane). No changes in expression were observed (n = 3). (**B**) Biotinylation assays of transfected cells for cell surface expression analysis. No changes in cell surface expression were observed (n = 3). Controls consisted of untransfected CHO cells. (**C**) Representative PIP strips of the phospholipid interaction of WT versus R359C (n = 3). 1 = Lysophosphatic acid, 2 = Lysophosphatidylcholine, 3 = Phosphatidylinositol, 4 = Phosphatidylinositol 3-phosphate, 5 = Phosphatidylinositol 4-phosphate, 6 = Phosphatidylinositol 5-phosphate, 7 = Phosphatidylethanolamine, 8 = Phosphatidylcholine, 9 = Sphingosine 1-phosphat, 10 = Phosphatidylinositol 3,4-bisphosphate, 11 = Phosphatidylinositol 3,5-bisphosphate, 12 = Phosphatidylinositol 4,5-bisphosphate, 13 = Phosphatidylinositol (3,4,5)-trisphosphate, 14 = Phosphatic acid, 15 = Phosphatidylserine, 16 = blank.

### THE p.Arg359Cys VARIANT ALTERS PI(4,5)P_2_ INTERACTION AND TRAPS CHANNELS IN THE PREOPEN-CLOSED STATE

p.Arg359 is homologous to p.Arg360 in K_v_7.1, which has been previously described as one of the key PI(4,5)P_2_ interaction sites^32,33^. The site of PI(4,5)P_2_-interaction is in proximity to a region formed by the cytoplasmic S6 and S5/S4-S5-linker regions in the open state (voltage-sensor module in upstate, S6-gate open)^29^ or closer to the S4/S4-S5-linker region in a preopen-closed state (voltage-sensor module in up-state, S6-gate closed)^28^. Both regions have been implicated in pharmacological effects of retigabine, which stabilizes open channel conformations of KCNQ channels^34^. Therefore, PI(4,5)P_2_ may interact dynamically with both regions depending on the S6-gate conformation. To investigate whether the dominant-negative LOF of the p.Arg359Cys variant in current density could be caused by a change in the interaction of K_V_7.5 channels with PI(4,5)P_2_, protein-phospholipid overlay assays were performed. The assays showed a significant reduction in the binding affinity of p.Arg359Cys channels compared to the WT to phosphatidylinositol (PI), phosphatidylinositol 5-phosphate (PI5P), (PI(3,4,5)P_3_), phosphatic acid, and most importantly, phosphatidylinositol 3,4-bisphosphate (PI(3,4)P_2_), phosphatidylinositol 4,5-bisphosphate (PI(4,5)P_2_), and phosphatidylinositol 3,5-bisphosphate (PI(3,5)P_2_; Fig 4C). Our results suggest that p.Arg359 plays an important role to confer binding to PI(4,5)P_2_ that is essential for channel opening, as Thomas et al. (2011) found for the homologous site in *KCNQ1*.

However, this experiment only yields answers on differences in selectivity and affinity of the subunit or channel to PI(4,5)P_2_, but does not deliver an explanation on a molecular level. It remains unclear in which conformational state of the channel this change in binding affinity does occur (open, closed or preopen-closed). Therefore, we performed consensus homology modeling (template structures: 2nz0.pdb, 3rbz.pdb, 5vms.pdb) combined with PI(4,5)P_2_-docking or direct homology modeling (template structure: 6V01.pdb) with PI(4,5)P_2_ present in the structure allowing us to generate two *KCNQ5* - PI(4,5)P_2_-models with alternative PI(4,5)P_2_-position depending on the state of the S6-gate (open vs. preopen-closed). Using these models, p.Arg359Cys could be introduced *in silico*. The residue 359 is positioned cytoplasmic to the S6-gate (Fig 5). Residue p.Arg359 contacts to residues p.Gln355, p.Gln356, p.His357, p.Gln360, p.Lys361, p.His362, p.Phe363 and p.Arg366 within the same subunit and residues p.Gln552, p.Ala555, p.Gly556 and p.Asn559 with the neighboring subunit (contact cutoff 4Å). When p.Arg359 is substituted by a cysteine, only the contact of p.C359 with p.Gln552 is conserved in the preopen-closed state model. On the contrary, in the open state model, p.Arg359 contacts to residues p.Gln355, p.Gln356, p.His357, p.His358, p.Gln360, p.Lys361, p.Pro534, and p.Tyr535 within the same subunit and residue p.Asp536 with the neighboring subunit (contact cutoff 4Å). The single contact to p.Asp536 of the neighboring subunits is lost for p.Cys359 *in silico*. According to this model, the p.Arg359Cys variant thus disrupts interactions with the neighboring subunit, particularly in the preopen-closed state model.

**Figure 5.**
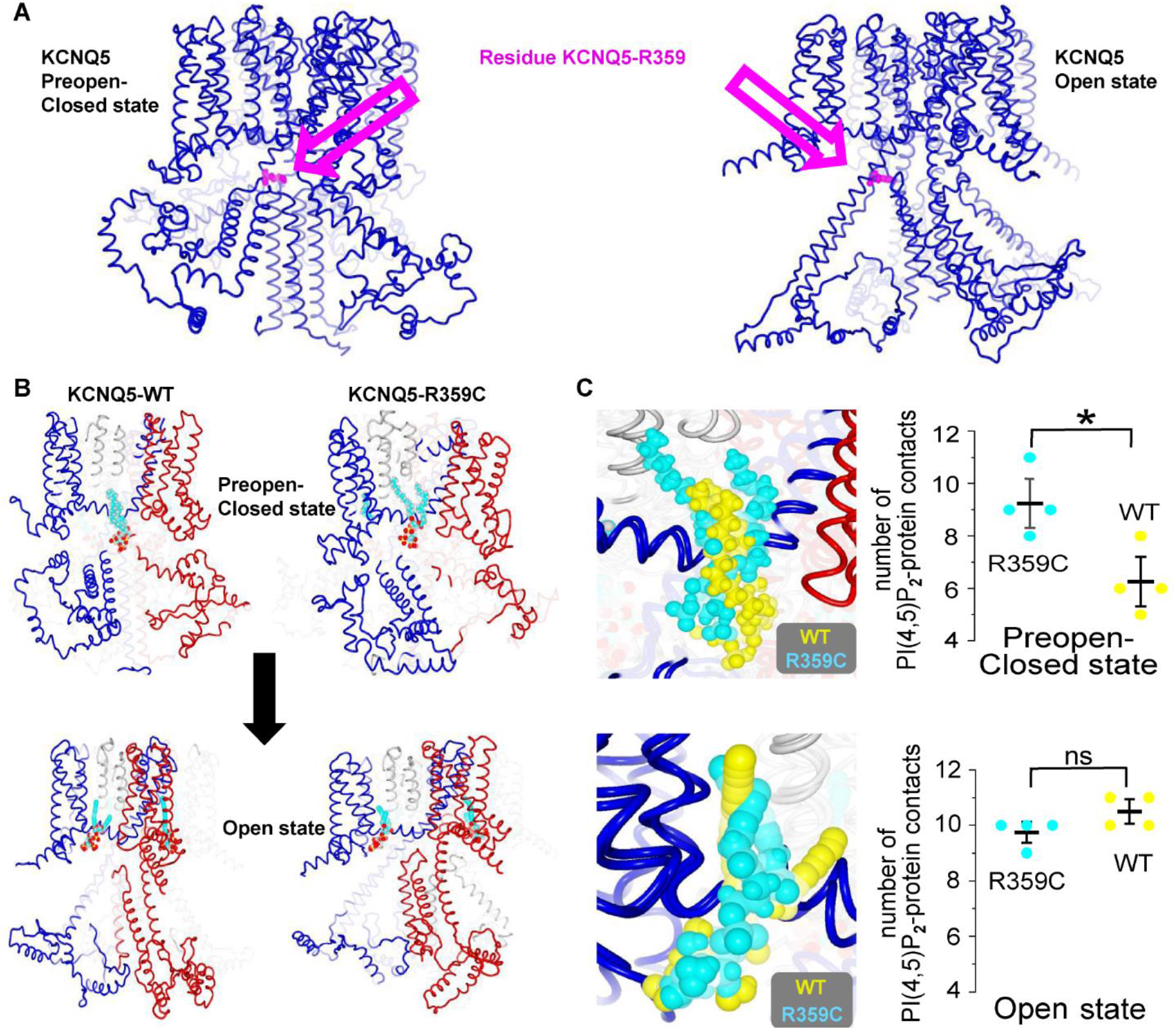
Model predictions for KCNQ5-WT and KCNQ5-R359C and their interaction with PI(4,5)P_2_. KCNQ5 preopen-closed state and open state models tethered with PI(4,5)P_2_ were generated as described in methods. Average structures over molecular dynamics simulations were calculated and are shown. (**A**) The two generated models are depicted with front-most arginine 359 (R359) shown in magenta and spacefill representation. (**B**) Wild Type (left) and R359C mutant (right) KCNQ5 preopen-closed state (upper figures) and open state models (lower figures) are shown. PI(4,5)P_2_ is shown in CPK-atom color code. Two adjacent KCNQ5-subunits are colored blue and red. (**C**) Section enlaged images show the position of PI(4,5)P_2_ in preopen-closed and open state models. The number of contacts are represented.

The interaction of PI(4,5)P_2_ with the respective K_V_7.5 WT or mutant model could be studied by molecular dynamics simulations. First, we simulated the impact of the disease-associated p.Arg359Cys variant. The open-state models were structurally stable over 3×32 nsec simulations (average root mean square deviations-RMSD of 11.88Å). Interestingly, in regions of PI(4,5)P_2_- binding the structures were even more stable suggesting that the lipid aids stability of the respective state (average RMSD: 5.32Å (PI(4,5)P_2_). In the preopen-closed state model with PI(4,5)P_2_ at the lower S6-S5/S4-S5-linker site PI(4,5)P_2_ was stably coordinated for 26 nsec (average RMSD of 6.9Å). Similar to the open-state model simulations the PI(4,5)P_2_-molecules the lipids were firmly bound to stabilize the state (average RMSD: 5,87Å). Therefore, the variant p.Arg359Cys did not interfere with *in silico* positioning of (PI(4,5)P_2_ to the K_V_7.5 models.

To estimate the interaction network of the PI(4,5)P_2_-molecules within their binding sites, contact analyses with a cutoff of 4Å were conducted. In the K_V_7.1-homologous open state models, the average number of PI(4,5)P_2_-K_V_7.5 interactions showed very similar contacts at 9.75±0.25 contacts per PIP_2_ molecule or 10.5±0.3 contacts per PIP_2_ molecule for K_V_7.5 WT vs. p.Arg359Cys (Student’s t-test, p=0.097). However, in the preopen-closed state K_V_7.5-models, the average PI(4,5)P_2_-K_V_7.5 is significantly different at 6.0±5.0 or 11.0±7.6 contacts per PIP_2_ per molecule for K_V_7.5 WT vs. p.Arg359Cys (Student’s t-test, p=0.04) (Fig 5). The number of contacts is an indicator of interaction energies, and less contact indicates reduced interaction energy. Our results suggest that p.Arg359Cys increases PI(4,5)P_2_-affinity to the preopen-closed state. On the contrary, this effect is not detected in the open state. As a result the preopen-closed state is relatively stabilized over the open channel state. The mutant channel is thus trapped by PI(4,5)P_2_ in its preopen-closed state thereby offering an explanation of reduced current density seen in the mutant channels *in vitro*.

## DISCUSSION

Here we analyzed clinical and genetic data from multiple cohorts of altogether 1292 independent families with GGE to identify and functionally characterize five likely disease-related variants suggesting that either haploinsufficiency or dominant-negative effects of *KCNQ5* are associated with GGE with or without mild to moderate ID. A hypothesis-driven statistical evaluation in three large GGE cohorts and matched controls indicated that rare functionally relevant variants in *KCNQ5* might be more frequent in GGE than expected by chance. Since *KCNQ5* was not among the top-ranking genes in the exome-wide primary analyses of these studies^20,21,23^, the statistical evidence remains only suggestive.

The phenotypes ranged from mild CAE to pharmaco-resistant early-onset absence epilepsy or JME with moderate ID. Most of the individuals (8/10) presented with absence seizures. In four individuals, developmental delay or ID was present prior to epilepsy. The phenotype comprising absence seizures and ID is in accordance with a previous report of an individual with mild ID and absence seizures in adolescence, carrying an intragenic duplication in *KCNQ5* leading to haploinsufficiency^17^. Lehman and colleagues reported four individuals suffering from ID and/or epilepsy caused by *KCNQ5* variants. However, the epilepsy type and ID extent of their individuals/cases differ from ours. Three of our variants were not associated with ID (p.Leu692Val, p.Gln735Arg and p. Ala301Glyfs*64). Moreover, one of the previously described individuals carried the first and only described GOF variant in *KCNQ5* (p.Pro369Arg), presenting the most severe phenotype published so far, with a severe DEE starting at the age of 5 months. None of our cases had a comparable severe phenotype and two carriers were even asymptomatic (one with p.Arg359Cys and one with p.Gln735Arg). The second most severely affected of the previously published individuals had a pharmaco-resistant epilepsy accompanied by severe ID. This individual had absence seizures too, but also focal seizures with impaired awareness and frontal epileptiform discharges in EEG (focal seizures have not been observed in any of the individuals reported by us). The remaining two individuals had mild to moderate ID without epilepsy. Interestingly, all four individuals from this report suffered from ataxia with a varying degree of severity^16^. In contrast, only one individual of our cohort, the most severely affected one, also had mild ataxia on neurological examination, the others did not have any neurological abnormalities on examination.

Five of ten individuals became seizure-free with or without medication and for one of ten individuals we did not obtain information about pharmaco-response. Lamotrigine seemed to be effective in carriers of p.Arg359Cys, and one of the carriers of this variant became seizure-free without medication. Valproate, topiramate, levetiracetam,ethosuximide and zonisamide were also prescribed but there is no indication that one of the drugs is more effective than the others.

The results of our functional analysis show a LOF in current density for homomeric channels for all investigated variants, and three of the investigated variants have a dominant-negative effect on current density in heteromeric expression experiments with a near complete loss-of-function for the p.Arg359Cys variant. The frameshift variant results in a premature stop codon in the pore region deleting the entire C-terminus. As the C-terminus comprises the interaction sites for the subunits to for a channel^35^, the absence of it causes the variant to be unable to form channels under homomeric expression and abolishes interaction with the WT subunits under heteromeric conditions causing a haploinsufficiency, but not a dominant-negative effect on the WT subunits. According to our electrophysiological studies, members of family 1 carried the most severe variant (p.Arg359Cys), leading to a severe LOF with a dominant-negative effect. We could not find clear genotype-phenotype correlations, since (i) family1 carrying the most severe variant displays a large phenotypic heterogeneity ranging from mildly to severely affected, pharmaco-resistant individuals, (ii) individuals in family 3 exhibited a similarly severe phenotype albeit the p.Gln735Arg variant showed a less prominent electrophysiological dysfunction, and (iii) also the phenotype of previously reported individuals carrying variants which only caused a haploinsufficiency were reported with similar or more severe phenotypes^16,17^. This is in contrast to other K_v_7 channels, in which functionally more severe variants with dominant-negative effects cause more severe epileptic phenotypes^36^. For *KCNQ5*, it rather seems that any significant LOF can contribute to an epileptic phenotype or ID of varying severity, which might be influenced by other individually differing factors, such as compensatory effects, the genetic background or environmental determinants. Larger cohorts are needed to further investigate this issue.

Remarkably, all missense variants showed stable total and membrane-expressed protein levels in CHO cells as compared to the WT. The LOF in current density is thus not caused by defects prior to membrane insertion of the channel, such as an abolished tetramerization or trafficking defect, as have been described for variants in *KCNQ2* and *KCNQ3*^36,37^. Rather, the three investigated missense variants have functional effects on channel gating, and thus, might mark important sites involved in channel opening. K_V_7 channels form the molecular basis of M-currents and are classically negatively regulated by muscarinic acetylcholine receptors via a PI(4,5)P_2_-dependent mechanism. PI(4,5)P_2_ is required for the stabilization of the open state relative to the closed state and PI(4,5)P_2_-depletion upon activation of muscarinic acetylcholine receptors leads to channel closure. Here, we provide evidence that the p.Arg359Cys variant causes altered PI(4,5)P_2_ binding and leads to stabilization of the mutant channels in the closed state, as according to our molecular modeling studies the interaction of PI(4,5)P_2_ with mutant channels is specifically enhanced in the preopen-closed and not in the open state.

Both other variants that we investigated are located in an evolutionary highly conserved region of the K_v_7.5 C-terminus which does not show any variants in unaffected individuals in multiple databases and is absent or not conserved in other K_v_7 channels, underlining the importance of this region for proper channel function in K_v_7.5. As the functional aspects of this region on channel behavior have not been described previously, these two variants might be able to elucidate binding partners and disclose the function of the distal C-terminus in channel opening. Investigating these molecular mechanisms may open doors for new treatment options in the future, especially for pharmaco-resistant patients.

In summary, we have identified rare loss-of-function variants in *KCNQ5* in five independent families, which are likely contributing to the pathophysiology of GGE. Two variants cause haploinsufficiency, three showed a dominant-negative effect on WT K_V_7.5 and K_V_7.3 channels. We were also able to identify the importance of p.Arg359 as crucial for PI(4,5)P_2_ interaction and channel opening. Consequently, the M-current in these individuals is likely reduced causing a decrease in action potential threshold and increased excitability of neurons expressing K_V_7.5 channels, thus leading to an elevated seizure susceptibility. The types of neurons and networks that are involved need to be determined in further studies.

## Data Availability

All data referred to in the manuscript is available upon request.

## Acknowledgement statement (including conflict of interest and funding sources)

We thank all the patients and their families for participating in this study. We also thank the following consortia for providing the variants and phenotypic data of the investigated individuals: EuroEPINOMICS-CoGIE Consortium (six individuals from two families), Epi25 Collaborative (one individual), and Epi4K (three individuals from three families).

This work was supported by the Research Unit FOR-2715, funded by the German Research Foundation (DFG) and the Fond Nationale de la Recherche (FNR) in Luxembourg (grants Le1030/16-1 to HL, INTER/DFG/17/11583046 to RK/PM, and We4896/4-1 to YGW), by the German Federal Ministry for Education and Research (BMBF, Treat-ION, 01GM1907 to HL, YGW, RK, PM) and by the European Science Foundation (EuroEPINOMICS-CoGIE project, grants from national funding agencies: DFG Le1030/11-1/2 to HL, FNR INTER/ESF/10/02/CoGIE to Rudi Balling/RK/PM). The foundation ‘no epilep’ funded patient recruitment (to HL). Epi25 was supported by the National Human Genome Research Institute (NHGRI) grants UM1 HG008895 and 5U01HG009088-02, and the Stanley Center for Psychiatric Research at the Broad Institute. The Epi4K study was supported by a National Institute of Neurological Disorders and Stroke (NINDS) National Institutes of Health (NIH) grant (ID: U01NS077367). We acknowledge the contribution of the HPC facilities of the University of Luxembourg (http://hpc.uni.lu) for computational support.

The authors report no competing interests.

## Authors contributions

JKr, JS, PM, GL, SM and HL designed the study and experiments. JKr, JS, AL, JH, MM, GS, PY, MK, MSH, PM performed experiments and analyzed data. JKr, JS, AL, JH, MM, GS, PY, MK, RK, PM, GL, SM, and HL, interpreted data. JKe, KA, HC, BJS, YGW, PK-K, SFB, GL, and HL recruited and phenotyped patients. JKr, JKe, GS, MK, PM, GL, SM, and HL wrote the manuscript. All authors read, revised and approved the manuscript.

